# Pembrolizumab monotherapy for previously treated metastatic HER2-negative breast cancer with germline *APOBEC3B* deletion: results of the phase II AUROR study

**DOI:** 10.1101/2024.08.07.24311537

**Authors:** Gwo Fuang Ho, Soo Chin Lee, Anita Zarina Bustam, Adlinda Alip, Nur Fadhlina Abdul Satar, Marniza Saad, Rozita Abdul Malik, Siew Eng Lim, Samuel G. W. Ow, Andrea Wong, Wan-Qin Chong, Yvonne L. E. Ang, Audrey Weng Yan Lee, Siti Norhidayu Hasan, Nabilah Tuan Zaid, Kian Boon Law, Yok Yong Toh, Hooi Chiao Tan, Bawani Selvam, Joanna Lim, Jia-Wern Pan, Soo Hwang Teo

## Abstract

**Background:** A common germline deletion polymorphism in the *APOBEC3B* gene increases the rate of somatic hypermutation in breast cancer, which in turn is associated with greater neoantigen burden and immune activation. This phase II study evaluated the impact of the *APOBEC3B* deletion polymorphism on the response to pembrolizumab monotherapy in metastatic HER2-negative breast cancer patients.

**Patients and methods:** Eligible patients had a confirmed diagnosis of metastatic HER2-negative breast cancer, 1-3 prior lines of therapy, and documented homozygous or heterozygous germline deletion of *APOBEC3B*. Patients received 200 mg of pembrolizumab intravenously every 3 weeks for up to 2 years. The primary endpoint was objective response rate. Secondary endpoints were disease control rate, progression-free survival, and overall survival.

**Results:** All enrolled patients (N = 44) were women, 36% had PD-L1-positive tumours, and 62% had received ≥2 previous lines of therapy for metastatic disease. ORR (95% CI) was 20.5% (9.8 – 35.5) in the total and 30.0% (6.7 – 65.3) in the PD-L1-positive populations. Disease control rate (95% CI) was 52.3% (36.7 – 67.5) and 40% (12.2 – 73.8), respectively. Median PFS was 3.1 months (95% CI, 2.1 – 4.3), and 6-month PFS rate was 29.5% (95% CI, 18.7 – 46.6). Median OS was 15.2 months (95% CI, 11.7 – 26.5), and 12-month OS rate was 60.2% (95% CI, 46.5 – 77.7). Treatment-related adverse events occurred in 30 (68.2%) patients, including 1 (2.3%) with grade 3 AE. There were no deaths due to AEs.

**Conclusions:** Pembrolizumab monotherapy demonstrated durable antitumour activity in a subset of previously treated metastatic HER2-breast cancer patients with germline APOBEC3B deletion.

**Clinical trial registration:** ClinicalTrials.gov, NCT03989089.

## Introduction

As standalone treatments or in combinations with conventional or targeted therapies, immunotherapy continues to redefine modern oncology. Despite this, only limited inroads have been made thus far in the treatment of breast cancer, the most commonly diagnosed cancer worldwide^1^. The current FDA approvals for immunotherapy in patients with breast cancer are: (1) pembrolizumab for patients who have the tumour agnostic biomarkers of microsatellite-instability high (MSI-H)^2,3^ or high tumour mutational burden (TMB >=10 mut/Mb)^4^; (2) pembrolizumab in combination with chemotherapy for metastatic TNBC in patients with PDL1 CPS ≥ 10^5^; and (3) pembrolizumab in combination with chemotherapy as neoadjuvant treatment in high-risk early stage TNBC^6^.

The identification of predictive biomarkers for immune checkpoint inhibitor (ICI) therapy remains a valuable endeavor as such biomarkers would enable better patient selection and clinical decision-making while reducing unnecessary financial costs and side effects for non-responders^7^. Exploratory analyses have tested settings and patients in whom immune checkpoint inhibitors may provide therapeutic benefit and these reveal that tumour PD-L1 expression may have some, albeit modest, predictive value in the metastatic TNBC setting; in addition, CD8, sTILs, and immune gene expression profiles may be associated with improved clinical outcomes of pembrolizumab^8,9^.

Response to ICI therapy has long been associated with high tumour mutational burden (TMB), which has been approved as a tumour-agnostic biomarker and is thought to be a surrogate for high neoantigen load leading to T-cell activation. In a recent pan-tumour retrospective analysis, which included TNBC samples, tumour mutational burden (TMB) ≥ 175 mutations/exome (mut/exome) was associated with improved antitumour activity of pembrolizumab^10^. A relative increase in tumour mutational burden (TMB) above the cohort mean (≥1.63 mutations/megabase [mut/Mb]) was observed in up to 36% of patients with mTNBC and conferred intrinsic survival benefits and possible susceptibility to anti–PD-1/PD-L1 monotherapy^11^. Data from the KEYNOTE-119 and KEYNOTE 86A studies demonstrated a potential positive association between TMB and response to pembrolizumab in patients with previously treated mTNBC^9,12^. Other biomarkers associated with response to anti–PD-1/PD-L1 therapy in patients with advanced solid tumours include high microsatellite instability^8^, mutations in BRCA1 or BRCA2, or both, and other genes involved in homologous recombination repair of double-stranded DNA breaks^13^.

One major source of TMB in breast cancer is APOBEC-associated DNA cytosine deaminase activity^14,15^. These enzymes are normally effector proteins in the innate immune response to viral infections, but upregulation of these enzymes during cancer development also causes elevated levels of genomic C-to-U deamination events, which can lead to a distinctive somatic hypermutation phenotype of cytosine mutation clusters (kataegis) in breast tumours^16,17^. Interestingly, a common germline deletion polymorphism in the *APOBEC3B* (*A3B*) gene occurs much more frequently in Asian and Oceanic women compared to Caucasian women and carriers of *A3B* germline deletion are more likely to develop cancer^18–20^. Breast cancers in *A3B* germline deletion carriers are more likely to have a C>T hypermutator phenotype^17^. These hypermutated cancers in turn have been shown to have a higher neoantigen burden and higher levels of immune activation^20–22^.

Taken together, these data suggest that *A3B* germline deletion carriers may have several features associated with benefit to pembrolizumab therapy (high TMB, higher immune activation, higher TILs). Given these considerations, we initiated a phase II, single arm, open label, Simon two-stage study of pembrolizumab in metastatic HER2-negative breast cancer patients (the AUROR trial) to evaluate the impact of the common germline deletion polymorphism in *APOBEC3B* on the response to pembrolizumab monotherapy.

## Methods

### Study design and patient population

AUROR (ClinicalTrials.gov, NCT03989089) was a phase II, single arm, open-label, Simon two-stage study of pembrolizumab monotherapy in metastatic HER2-negative breast cancer patients with germline *APOBEC3B* deletion. Using DNA extracted from peripheral blood, germline APOBEC3B polymorphism status was determined using single tube PCR assay with one forward (5’CCTGTCCCTTTTCAGAATTTAAGC-3’) and two reverse primers flanking the deletion (5’-CTTGATCGGGAGCATAC-3’, 5’TGGAGCCAATTAATCACTTCAT-3’; Klonowska et al. 2017^23^). In total, 146 patients were screened, and 92 patients (63%) were found to be either homozygous or heterozygous carriers. 44 adult patients eligible for enrolment had histologically confirmed HER2-negative breast cancer (infiltrating ductal or lobular breast carcinoma) with measurable metastatic disease based on RECIST 1.1; received at least one but not more than three (3) prior lines of palliative chemotherapy for metastatic breast cancer; received at least one line of hormonal therapy in the metastatic setting for patients with ER-positive breast cancer; documented germline *APOBEC3B* mutation; an ability to provide archival tumour tissue sample or tissues from a newly obtained core or excisional biopsy of a tumour lesion not previously irradiated; an Eastern Cooperative Oncology Group performance-status score of 0 or 1; a life expectancy of at least 3 months; and adequate organ function. Exclusion criteria were the use of any investigational agent or participation in another therapeutic clinical trial concurrently or in the 30 days prior to inclusion; previous therapy that targeted PD-1, PD-L1, PD-L2, or an agent directed at another coinhibitory T-cell receptor; active autoimmune disease warranting systemic treatment within the previous 2 years; immunosuppressive therapy or diagnosis of immunodeficiency within the previous 1 week; active central nervous system metastases or carcinomatous meningitis (previously treated stable brain metastases were permitted); a history of non-infectious pneumonitis requiring treatment with steroids or current pneumonitis; or any active infection warranting systemic therapy.

### Study Intervention

Eligible patients received single agent pembrolizumab, administered as 30-minute intravenous infusions every 3 weeks for up to two years; or until confirmed disease progression or unacceptable toxic effects had occurred or withdrawal of consent or physician’s decision. Following evidence of radiological PD by RECIST 1.1 criteria, clinically stable patients who continued to achieve a clinically meaningful benefit were allowed to remain on treatment at the physician’s discretion.

### Assessments

Response was assessed by imaging every 9 weeks in the first year, then every 12 weeks thereafter using RECIST, version 1.1, by blinded independent central review. After verified disease progression or initiation of new anticancer therapy, patients were monitored for survival status every 12 weeks. Baseline PD-L1 expression in archival or newly obtained formalin-fixed tumour samples was assessed at a central laboratory with the use of PD-L1 IHC 22C3 pharmDx (DAKO) and characterized according to the Combined Positive Score (CPS). Adverse events were monitored throughout the trial and for 30 days (90 days for serious adverse events) after discontinuation of the trial regimen and graded according to the National Cancer Institute Common Terminology Criteria for Adverse Events, version 4.0. Immune-mediated adverse events were determined based on a prespecified list of Medical Dictionary for Regulatory Activities (MedDRA) terms, which was updated with each new version of MedDRA.

### Endpoints

The primary endpoint for this study was objective response rate (ORR), defined as the proportion of patients with complete response (CR) or partial response (PR) assessed using RECIST v1.1 by an independent central radiology review. Patients who passed away prior to the first disease assessment were considered to have progressive disease (clinical disease progression) as their best response. Secondary endpoints included disease control rate [DCR; defined as the proportion of patients with CR, PR or stable disease (SD) at 6, 12, and 24 months based on RECIST v1.1], duration of response [DOR; defined as the time from documentation of tumour response to disease progression for subjects who demonstrate CR, PR or SD], progression-free survival [PFS; defined as the time from start of study medication until the first sign of disease progression based on RECIST v1.1 or death from any cause, whichever occurred first], and overall survival [OS; defined as the date from study commencement to the date of death from any cause, censoring patients who were still alive at data cut-off].

### Trial Oversight

The trial was developed by the study team. The trial protocol and all amendments were approved by the appropriate ethics committee at each participating institution. All patients provided written informed consent before enrolment.

### Statistical analysis

Efficacy and safety were assessed in the intention-to-treat population. 95% confidence interval (CI) for proportion was calculated using Clopper-Pearson exact method. The Kaplan–Meier (KM) method was used to estimate progression free survival (PFS) and overall survival (OS). Median survival and exact 95% CI were calculated. Statistical analysis was completed using R version 4.3.1.

## Results

### Patient characteristics

Between July 2020 and June 2023, 159 patients were screened for eligibility. Of these patients, 44 patients passed all screening procedures and were recruited to the study. Out of the 115 patients who did not pass screening, approximately half [n = 55] failed screening due to the absence of germline *APOBEC3B* deletion, while the remaining 60 patients did not meet the eligibility criteria. Of the 44 enrolled patients, 10 had homozygous germline *APOBEC3B* deletion, while 34 were heterozygous carriers of the deletion.

For the enrolled patients, the median duration of treatment was 4.2 months (range, 0.6-43.0), during which patients received a median of 6 cycles of pembrolizumab (range, 1-35) with a median duration of pembrolizumab exposure of 125 days (range, 18-743). Forty (90.9%) patients had discontinued pembrolizumab as of the data cut-off primarily due to disease progression [n = 38 (86.4%)].

All enrolled patients were women, with a median age of 58 years (range, 32-83). Twenty-three (52%) patients had histologically confirmed metastatic HR+/HER2-disease, while 20 (45%) had metastatic triple-negative breast cancer (TNBC). Of the 44 patients, 16 (36%) had 1 prior line of palliative chemotherapy for metastatic disease, 14 (32%) had 2 prior lines and 13 (30%) had 3 or more prior lines (Table 1).

**Table 1.** Clinico-pathological characteristics of the AUROR trial patients.

| Characteristic |  | Median/N | Range/% |
| --- | --- | --- | --- |
| Age, years, median (range) |  | 58 | 32-83 |
| ECOG performance status, *n* (%) | 0 | 31 | 70% |
|  | 1 | 13 | 30% |
| ER status, *n* (%) | Positive | 23 | 52% |
|  | Negative | 20 | 45% |
|  | NA | 1 | 2% |
| Previous lines of chemotherapy, *n* (%) | 1 | 16 | 36% |
|  | 2 | 14 | 32% |
|  | 3 | 13 | 30% |
|  | NA | 1 | 2% |
| Previous lines of hormonal therapy, *n* (%) [n=23 ER+] | 1 | 5 | 22% |
|  | 2 | 10 | 43% |
|  | 3 or more | 8 | 35% |
| Previous lines of targeted therapy, *n* (%) | 0 | 31 | 70% |
|  | 1 | 10 | 23% |
|  | 2 | 2 | 5% |
|  | NA | 1 | 2% |
| PD-L1 status, *n* (%) | Positive | 12 | 27% |
|  | Negative | 28 | 64% |
|  | NA | 4 | 9% |
| *APOBEC3B* germline status, *n* (%) | WT/Del | 33 | 75% |
|  | Del/Del | 11 | 25% |

### Treatment efficacy

Of the 44 enrolled patients, 1 (2.3%) patient had a CR and 8 (18.2%) patients had a PR, for an ORR of 20.5% (95% CI, 9.8 – 35.3, Figure 1, Supp. Table 1). An additional 14 (31.8%) patients had SD, resulting in a DCR of 52.3% (95% CI, 36.7 – 67.5). The DCR at 6 months was 25.0% (95% CI, 13.2 – 40.3). The median duration of response (DOR) was 4.1 months (range, 0.8 – 37.5).

**Figure 1.**
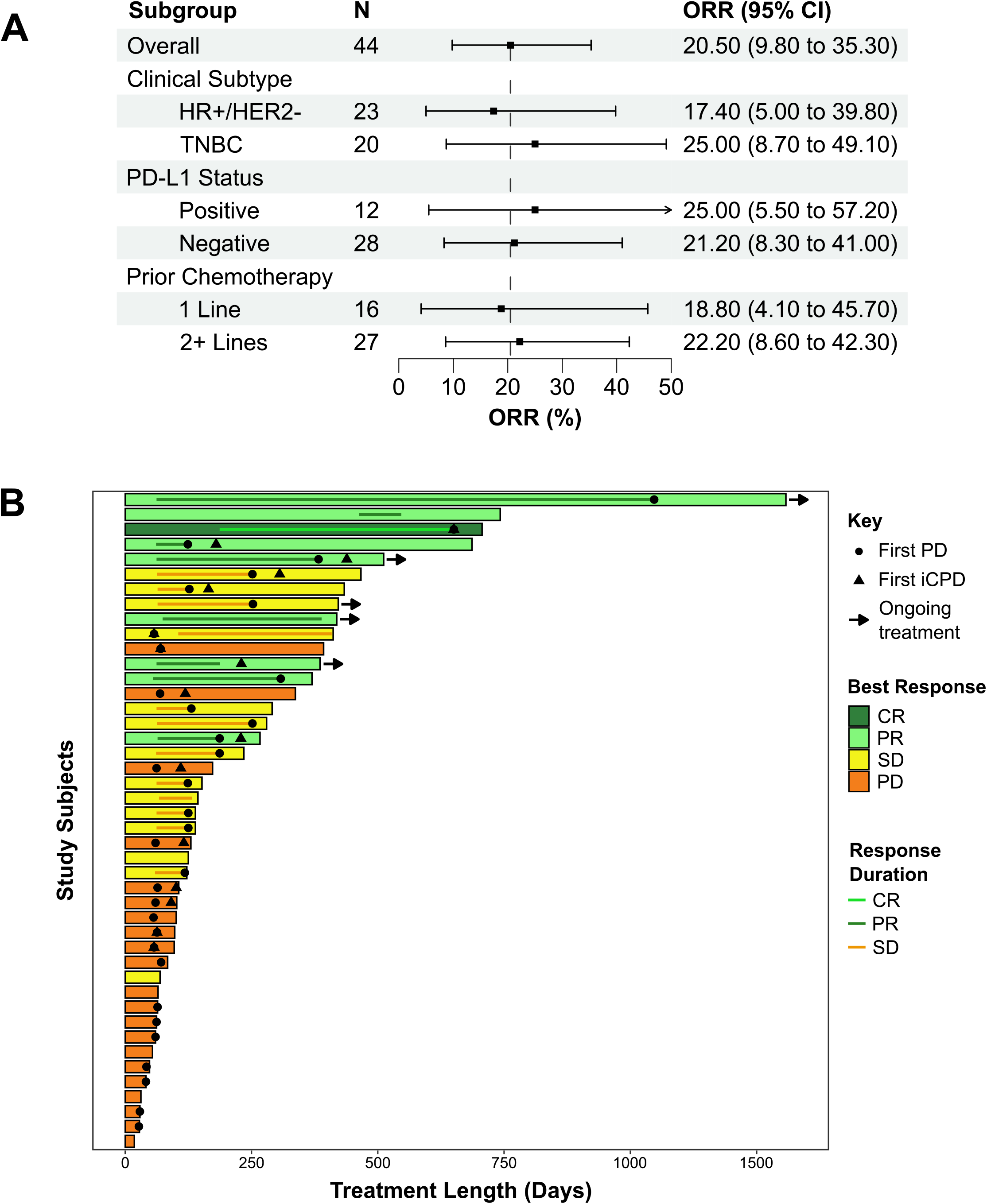
Efficacy of treatment in the AUROR trial patients. **(A)** Objective response rate as assessed by RECIST v1.1 in the overall cohort and subgroups of the efficacy population (N = 44). All specified subgroups are exploratory. **(B)** Time to response and response duration as assessed by RECIST v1.1. CR, complete response; PD, progressive disease; PR, partial response; SD, stable disease.

At the time of data cut-off, 39 (88.6%) patients had disease progression or had passed away. Median PFS was 3.1 months (95% CI, 2.1 – 4.3, Supp. Fig. 1), with 6- and 12-month PFS rates of 29.5% (95% CI, 18.7 – 46.6) and 17.5% (95% CI, 9.1 – 33.8), respectively. By data cut-off, 31 (70.5%) patients had passed away. Median OS was 15.2 months (95% CI, 11.7 – 26.5, Supp. Fig. 1) with 6- and 12-month OS rates of 81.8% (95% CI, 71.2 – 94.0) and 60.2% (95% CI, 46.5 – 77.7), respectively.

Subgroup analyses (Figure 1, Supp. Fig 2-4, Supp. Table 2-4) indicated that, of the 23 HR+/HER2-patients, 1 had a CR and 3 had a PR, for an ORR of 17.4% (95% CI, 5.0 – 38.8), with a median DOR of 2.1 months. Of the 20 patients with TNBC, 5 had a PR, for an ORR of 25.0% (95% CI, 8.7 – 49.1) and a median DOR of 6.2 months. 3 out of 12 patients with PD-L1-positive disease (CPS ≥ 10) had a PR [ORR = 25.0% (95% CI, 5.5 – 57.2), median DOR = 5.2 months], while 6 out of 28 patients with PD-L1-negative disease (CPS < 10) had a PR [ORR = 21.2% (95% CI, 8.3 – 41.0), median DOR = 3.2 months]. The ORR for patients with 1 prior line of chemotherapy was 18.8% (95% CI, 4.1 – 45.7; 3/16 patients with PR; median DOR = 2.1 months) while the ORR patients with 2 or more prior lines of chemotherapy was 22.2% (95% CI, 8.6 – 42.3; 6/27 patients with CR or PR; median DOR = 6.3 months). Additionally, 7/34 (20.6%; median DOR = 5.2 months) patients who were heterozygous for the A3B deletion had an objective response, compared to 2/10 (20.0%; median DOR = 4.0 months) patients who were homozygous for the A3B deletion.

### Safety

Of the 44 enrolled patients, all were evaluable for safety. Thirty (68.2%) patients experienced one or more treatment-related AEs of any grade. The most common treatment-related AEs were skin rash (20.5%) and hyperthyroidism (15.9%). Only 1 (2.3%) patient had immune-mediated grade 3 AE, a case of grade 3 immunotherapy-related colitis. The safety profile of pembrolizumab therapy in this study was consistent with that observed in previously reported studies, and no new safety signals were identified.

## Discussion

The results of the protocol-specified analysis showed that single-agent pembrolizumab resulted in objective response rate of 20.5% in germline carriers of the *APOBEC3B* deletion polymorphism. Responses to pembrolizumab were durable over longer follow-up.

Our patient cohort included an almost equal amount of ER+/HER2- (53%) and TNBC (47%) patients who were heavily pre-treated, with 62% of patients having received 2 or more previous lines of palliative chemotherapy. In this setting, our ORR of 20.5% (and median OS of 15.2 months) for pembrolizumab monotherapy is noteworthy. Prior studies of pembrolizumab monotherapy in previously treated metastatic TNBC have reported much lower ORRs, with KEYNOTE-086A reporting an overall ORR of 5.3% (median OS of 9.0 months)^24^ and KEYNOTE-119^25^ reporting an overall ORR of 9.6% (median OS of 9.9 months). Similarly, a phase I study of treatment with single-agent atezolizumab (anti-PD-L1 monoclonal antibody) in second-line or greater metastatic TNBC patients reported an ORR of only 6.4%^26^.

The response rate in our ER+/HER2- subgroup was 17.4%, which was numerically lower but not statistically different from our TNBC subgroup (25% ORR). This response rate is also numerically higher than previous reports for pembrolizumab monotherapy in this patient subgroup - in the phase Ib KEYNOTE-028 trial, pembrolizumab was tested as a single agent in 25 patients with heavily pretreated HR+/HER2- PD-L1 CPS >1 metastatic breast cancer^27^, and the authors reported an ORR of 12.0% (95% CI, 2.5 to 31.2) with a median duration of response of 12 months. Additionally, a phase II study of pembrolizumab in combination with metronomic cyclophosphamide in 20 metastatic endocrine-resistant breast cancer patients reported a response rate of 10.0%^28^. Thus, our results provide a strong rationale for further investigation of APOBEC gene variants as biomarkers for immunotherapy in breast cancer in both ER+/HER2- and TNBC patients.

In our study, PD-L1-positive patients (CPS ≥ 10) had a numerically higher ORR relative to our intention-to-treat population (25.0% vs 20.5%, respectively). This modest increase is in line with previous studies – KEYNOTE-355^5^ reported an ORR of 52.7% in patients receiving combination pembrolizumab-chemotherapy with PD-L1 CPS ≥ 10 compared to an ORR of 44.9% in patients with PD-L1 CPS ≥ 1. Similarly, KEYNOTE-119^25^ reported an ORR of 17.7% in patients with PD-L1 CPS ≥ 10 versus an ORR of 9.6% in the intent-to-treat PD-L1 unselected population. KEYNOTE-86A^24^, on the other hand, reported only a small numerical increase in ORR for PD-L1-positive patients compared to the total population (5.7% versus 5.3%, respectively). Thus, our study provides some support for the use of PD-L1 and *A3B* deletion testing in tandem to select patients most likely to respond to pembrolizumab therapy.

Our study had several limitations. The sample size for the study is relatively small, with a small absolute number of responders, which limits the power of our subgroup analyses. Our results are also not generalizable to patients who receive pembrolizumab in combination with chemotherapy, patients with less advanced disease, or patients with previously untreated metastatic breast cancer. Additionally, this study was conducted fully in Asian patients that are known to have a more active tumour immune microenvironment than their Western counterpart^29,30^, and the ORR seen in this study may have been influenced by population-specific effects.

In conclusion, the *APOBEC3B* deletion polymorphism may be a useful biomarker to select subsets of patients that are more likely to respond to pembrolizumab therapy. Further randomized studies of the *APOBEC3B* deletion polymorphism and other APOBEC variants as biomarkers for immunotherapy in breast cancer are warranted.

## Data Availability

All data produced in the present study are available upon reasonable request to the authors.

## Acknowledgements

The sponsor of this clinical trial was Cancer Research Malaysia. Cancer Research Malaysia receives charitable funding from the Khind Starfish Foundation, Estée Lauder Companies, Yayasan PETRONAS, and Yayasan Sime Darby which contributed to the funding of this study. Merck Sharp & Dohme (MSD) provided free access to the trial drug through the Merck Investigator Studies Program.

## Disclosures

GFH reports research grants as well as speaker, advisor, and training engagements with MSD. SCL reports grant support/research collaborations from Pfizer, Eisai, Taiho, ACT Genomics, Bayer, MSD, and Adagene as well as advisory board/speaker invitations from Pfizer, Novartis, Astra Zeneca, ACT Genomics, Eli Lilly, MSD, Roche, DKSH, and Daiichi Sankyo. NA reports research grants and speaking engagements with MSD. RAM reports consultancy engagements with MSD. AW reports advisory board/honorarium from Pfizer, Astra Zeneca, Eisai, Daiichi Sankyo, Novartis as well as research funding from Otsuka Pharmaceuticals. CWQ reports advisory board/speaker invitations from Merck, MSD, DKSH, Ipsen.

## Author Contributions

GFH was the principal investigator for this study, while SCL and SHT were co-PIs. GFH and SCL were the clinical leads and contributed to patient recruitment, study design, patient care and treatment, and project supervision. AZB, AA, FAS, MS, RAM, SEL, SO, AW, WQC, KS, and YA contributed to patient recruitment and patient care. YYT and HCT were the on-site study coordinators and contributed to patient recruitment and data collection. AWYL and SNH contributed to data collection and sample processing. KBL provided statistical support and contributed to data analysis and drafting of the manuscript and figures. BS and JL contributed to data collection, data analysis, and project supervision. JWP contributed to project direction and supervision, data analysis and drafting of the manuscript and figures. SHT also contributed to study design and project funding, drafted the manuscript, and contributed to project direction and supervision. The work reported in the paper has been performed by the authors, unless clearly specified in the text.

## Ethics Statement

Ethical approval for this work was given by the Medical Research Ethics Committee of the University of Malaya Medical Centre (ID: NMRR-18-3343-45490) and by the National Healthcare Group Domain Specific Review Board of the National University Hospital of Singapore (ID: 2019/00871). The study was conducted according to the principles of the Declaration of Helsinki, complied with all applicable regulations, was registered on ClinicalTrials.gov (NCT03989089).

## Notes

### Clinical Trial

NCT03989089

### Author Declarations

Medical Research Ethics Committee of the University of Malaya Medical Centre gave ethical approval for this work (ID: NMRR-18-3343-45490). National Healthcare Group Domain Specific Review Board of the National University Hospital of Singapore also gave ethical approval for this work (ID: 2019/00871).

## References

1. Bray Bsc, F. et al. Global cancer statistics 2022: GLOBOCAN estimates of incidence and mortality worldwide for 36 cancers in 185 countries. CA. Cancer J. Clin. 74, 229–263 (2024).

2. Marabelle, A. et al. Efficacy of pembrolizumab in patients with noncolorectal high microsatellite instability/mismatch repair–deficient cancer: Results from the phase II KEYNOTE-158 study. J. Clin. Oncol. (2020) doi:10.1200/JCO.19.02105.

3. Marcus, L., Lemery, S. J., Keegan, P. & Pazdur, R. FDA approval summary: Pembrolizumab for the treatment of microsatellite instability-high solid tumors. Clin. Cancer Res. (2019) doi:10.1158/1078-0432.CCR-18-4070.

4. Marcus, L. et al. FDA approval summary: Pembrolizumab for the treatment of tumor mutational burden-high solid tumors. Clin. Cancer Res. (2021) doi:10.1158/1078-0432.CCR-21-0327.

5. Cortes, J. et al. Pembrolizumab plus chemotherapy versus placebo plus chemotherapy for previously untreated locally recurrent inoperable or metastatic triple-negative breast cancer (KEYNOTE-355): a randomised, placebo-controlled, double-blind, phase 3 clinical trial. Lancet (2020) doi:10.1016/S0140-6736(20)32531-9.

6. Schmid, P. et al. Pembrolizumab for Early Triple-Negative Breast Cancer. N. Engl. J. Med. (2020) doi:10.1056/nejmoa1910549.

7. Elliott, M. J., Wilson, B. & Cescon, D. W. Current Treatment and Future Trends of Immunotherapy in Breast Cancer. Curr. Cancer Drug Targets (2022) doi:10.2174/1568009622666220317091723.

8. Cocco, S. et al. Biomarkers in triple-negative breast cancer: State-of-the-art and future perspectives. International Journal of Molecular Sciences 10.3390/ijms21134579 (2020).

9. Loi, S. et al. Association Between Biomarkers and Clinical Outcomes of Pembrolizumab Monotherapy in Patients With Metastatic Triple-Negative Breast Cancer: KEYNOTE-086 Exploratory Analysis. JCO Precis. Oncol. (2023) doi:10.1200/po.22.00317.

10. Cristescu, R. et al. Tumor mutational burden predicts the efficacy of pembrolizumab monotherapy: A pan-tumor retrospective analysis of participants with advanced solid tumors. J. Immunother. Cancer (2022) doi:10.1136/jitc-2021-003091.

11. Thomas, A. et al. Tumor mutational burden is a determinant of immune-mediated survival in breast cancer. Oncoimmunology (2018) doi:10.1080/2162402X.2018.1490854.

12. Winer, E. P. et al. Association of tumor mutational burden (TMB) and clinical outcomes with pembrolizumab (pembro) versus chemotherapy (chemo) in patients with metastatic triple-negative breast cancer (mTNBC) from KEYNOTE-119. J. Clin. Oncol. (2020) doi:10.1200/jco.2020.38.15_suppl.1013.

13. Cindy Yang, S. Y. et al. Pan-cancer analysis of longitudinal metastatic tumors reveals genomic alterations and immune landscape dynamics associated with pembrolizumab sensitivity. Nat. Commun. (2021) doi:10.1038/s41467-021-25432-7.

14. Burns, M. B., Temiz, N. A. & Harris, R. S. Evidence for APOBEC3B mutagenesis in multiple human cancers. Nat. Genet. (2013) doi:10.1038/ng.2701.

15. Roberts, S. A. et al. An APOBEC cytidine deaminase mutagenesis pattern is widespread in human cancers. Nat. Genet. 45, 970–976 (2013).

16. Nik-Zainal, S. et al. Mutational processes molding the genomes of 21 breast cancers. Cell 149, 979–93 (2012).

17. Nik-Zainal, S. et al. Association of a germline copy number polymorphism of APOBEC3A and APOBEC3B with burden of putative APOBEC-dependent mutations in breast cancer. Nat. Genet. 46, 487–491 (2014).

18. Long, J. et al. A common deletion in the APOBEC3 genes and breast cancer risk. J. Natl. Cancer Inst. 105, 573–579 (2013).

19. Xuan, D. et al. APOBEC3 deletion polymorphism is associated with breast cancer risk among women of European ancestry. Carcinogenesis (2013) doi:10.1093/carcin/bgt185.

20. Wen, W. X. et al. Germline APOBEC3B deletion is associated with breast cancer risk in an Asian multi-ethnic cohort and with immune cell presentation. Breast Cancer Res. 18, 56 (2016).

21. Cescon, D. W., Haibe-Kains, B. & Mak, T. W. APOBEC3B expression in breast cancer reflects cellular proliferation, while a deletion polymorphism is associated with immune activation. Proc. Natl. Acad. Sci. U. S. A. 112, 2841–2846 (2015).

22. Pan, J. et al. Germline APOBEC3B deletion increases somatic hypermutation in Asian breast cancer that is associated with Her2 subtype, PIK3CA mutations and immune activation. Int. J. Cancer ijc.33463 (2021) doi:10.1002/ijc.33463.

23. Klonowska, K. et al. The 30 kb deletion in the APOBEC3 cluster decreases APOBEC3A and APOBEC3B expression and creates a transcriptionally active hybrid gene but does not associate with breast cancer in the European population. Oncotarget 8, 76357–76374 (2017).

24. Adams, S. et al. Pembrolizumab monotherapy for previously treated metastatic triple-negative breast cancer: Cohort A of the phase II KEYNOTE-086 study. Ann. Oncol. (2019) doi:10.1093/annonc/mdy517.

25. Winer, E. P. et al. Pembrolizumab versus investigator-choice chemotherapy for metastatic triple-negative breast cancer (KEYNOTE-119): a randomised, open-label, phase 3 trial. Lancet Oncol. (2021) doi:10.1016/S1470-2045(20)30754-3.

26. Emens, L. A. et al. Long-term Clinical Outcomes and Biomarker Analyses of Atezolizumab Therapy for Patients with Metastatic Triple-Negative Breast Cancer: A Phase 1 Study. JAMA Oncol. (2019) doi:10.1001/jamaoncol.2018.4224.

27. Rugo, H. S. et al. Safety and antitumor activity of pembrolizumab in patients with estrogen receptor–positive/human epidermal growth factor receptor 2–negative advanced breast cancer. Clin. Cancer Res. (2018) doi:10.1158/1078-0432.CCR-17-3452.

28. Mery, B. et al. Pembrolizumab in Lymphopenic Metastatic Breast Cancer Patients Treated with Metronomic Cyclophosphamide: A Clinical and Translational Prospective Study. Breast Cancer Targets Ther. (2023) doi:10.2147/BCTT.S400055.

29. Kan, Z. et al. Multi-omics profiling of younger Asian breast cancers reveals distinctive molecular signatures. Nat. Commun. 9, 1725 (2018).

30. Pan, J. W. et al. The molecular landscape of Asian breast cancers reveals clinically relevant population-specific differences. Nat. Commun. 11, 1–12 (2020).

