## Supplemental Tables 1-4, Supplemental Figures 1-4 for "Pembrolizumab monotherapy for previously treated metastatic HER2-negative breast cancer with germline *APOBEC3B* deletion: results of the phase II AUROR study"

Supp. Table 1. Summary of efficacy

| | n (%) | 95% CI[a] |
|---|---|---|
| Best overall response | | |
| Complete response (CR) | 1 (2.3) | 0.0 – 12.0 |
| Partial response (PR) | 8 (18.2) | 8.2 – 32.7 |
| Stable disease (SD) | 14 (31.8) | 18.6 – 47.6 |
| Progressive disease (PD) | 17 (38.6) | 24.4 – 54.5 |
| Not evaluated | 4 (9.1) | 2.5 – 21.7 |
| ORR (CR+PR) | 9 (20.5) | 9.8 – 35.3 |
| DCR (CR+PR+SD) | 23 (52.3) | 36.7 – 67.5 |
| 6-month | 11 (25.0) | 13.2 – 40.3 |
| Median DOR months (range) | 4.1 (0.8 – 37.5) | |
| PFS, median months (95% CI) | 3.1 (2.1 – 4.3) | |
| 6-month | 29.5 (18.7 – 46.6) | |
| 12-month | 17.5 (9.1 – 33.8) | |
| OS, median months (95% CI) | 15.2 (11.7 – 26.5) | |
| 6-month | 81.8 (71.2 – 94.0) | |
| 12-month | 60.2 (46.5 – 77.7) | |

CI = confidence interval, DCR = disease control rate, ORR = overall response rate, OS = overall survival, PFS = progression free survival

[a]CI based on normal approximation to binomial estimation

Supp. Table 2. ORR, DCR and Median DOR according to ER status.

| | ER Positive (N=23) | | ER Negative (N=20) | |
|---|---|---|---|---|
| | n (%) | 95% CI[a] | n (%) | 95% CI[a] |
| ORR (CR+PR) | 4 (17.4) | 5.0 – 38.8 | 5 (25.0) | 8.7 – 49.1 |
| DCR (CR+PR+SD) | 12 (52.2) | 30.6 – 73.2 | 11 (55.0) | 31.5 – 76.9 |
| Median DOR months (range) | 2.1 (0.8 – 22.3) | | 6.2 (2.1 – 37.5) | |
| PFS, median months (95% CI) | 2.3 (2.0, 4.2) | | 4.2 (2.1, NA) | |
| OS, median months (95% CI) | 15.8 (13.0, 29.2) | | 11.1 (9.6, NA) | |

CI = confidence interval, DCR = disease control rate, ORR = overall response rate
[a]CI based on normal approximation to binomial estimation

Supp. Table 3. ORR, DCR and Median DOR according to PD-L1 status.

| | PD-L1-positive (N=12) | | PD-L1-negative (N=28) | |
|---|---|---|---|---|
| | n (%) | 95% CI[a] | n (%) | 95% CI[a] |
| ORR (CR+PR) | 3 (25.0) | 5.5 – 57.2 | 6 (21.2) | 8.3 – 41.0 |
| DCR (CR+PR+SD) | 4 (33.3) | 9.9 – 65.1 | 19 (67.9) | 47.6 – 84.1 |
| Median DOR months (range) | 5.2 (4.0 – 11.3) | | 3.2 (1.9 – 32.4) | |
| PFS, median months (95% CI) | 5.1 (2.10, NA) | | 4.1 (3.88, NA) | |
| OS, median months (95% CI) | 15.2 (11.7, NA) | | 18.6 (13.0, 29.9) | |

CI = confidence interval, DCR = disease control rate, ORR = overall response rate
[a]CI based on normal approximation to binomial estimation

Supp. Table 4. ORR, DCR and Median DOR according to number of prior lines of therapy.

| | 1 prior line (N=16) | | Two or more prior lines (N=27) | |
|---|---|---|---|---|
| | n (%) | 95% CI[a] | n (%) | 95% CI[a] |
| ORR (CR+PR) | 3 (18.8) | 4.1 – 45.7 | 6 (22.2) | 8.6 – 42.3 |
| DCR (CR+PR+SD) | 11 (68.8) | 41.3 – 89.0 | 12 (44.4) | 25.5 – 64.7 |
| Median DOR months (range) | 2.1 (0.8 – 22.3) | | 6.3 (2.1 – 37.5) | |
| PFS, median months (95% CI) | 4.0 (2.3, NA) | | 2.3 (2.0, 8.3) | |
| OS, median months (95% CI) | 13.0 (10.0, NA) | | 15.4 (11.7, 37.1) | |

CI = confidence interval, DCR = disease control rate, ORR = overall response rate
[a]CI based on normal approximation to binomial estimation

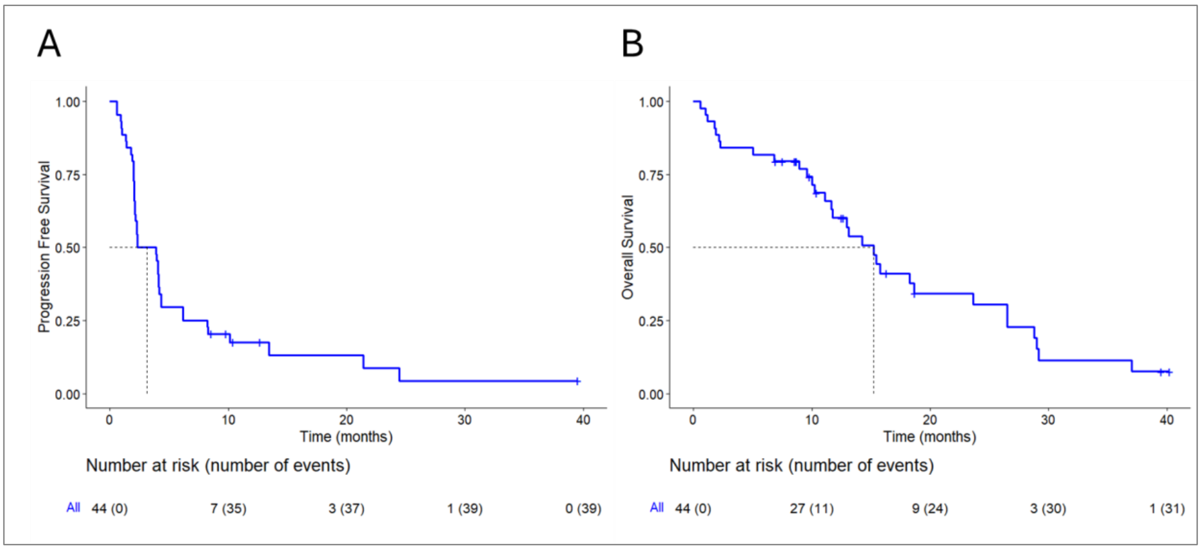


Supp. Fig. 1. Kaplan-Meier estimates of (A) progression free survival and (B) overall survival.

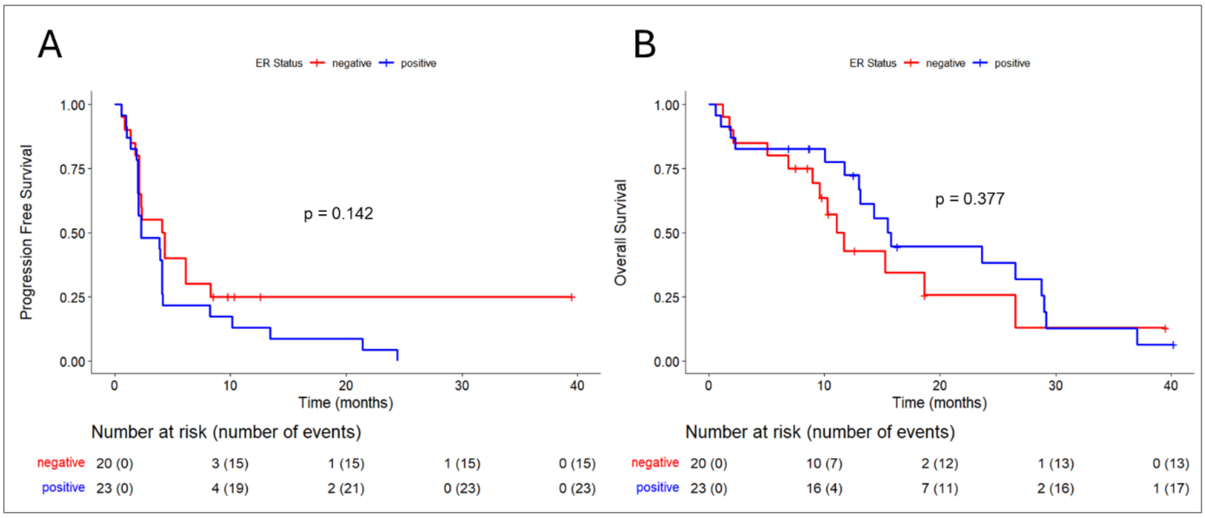


Supp. Fig 2. Kaplan-Meier estimates of (A) progression free survival and (B) overall survival according to ER status.

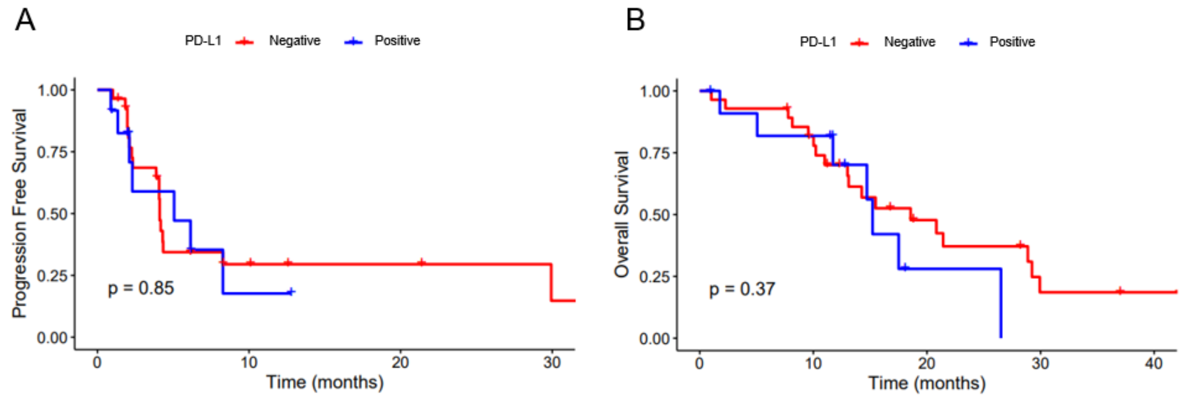


Supp. Fig 3. Kaplan-Meier estimates of (A) progression free survival and (B) overall survival according to PD-L1 status.

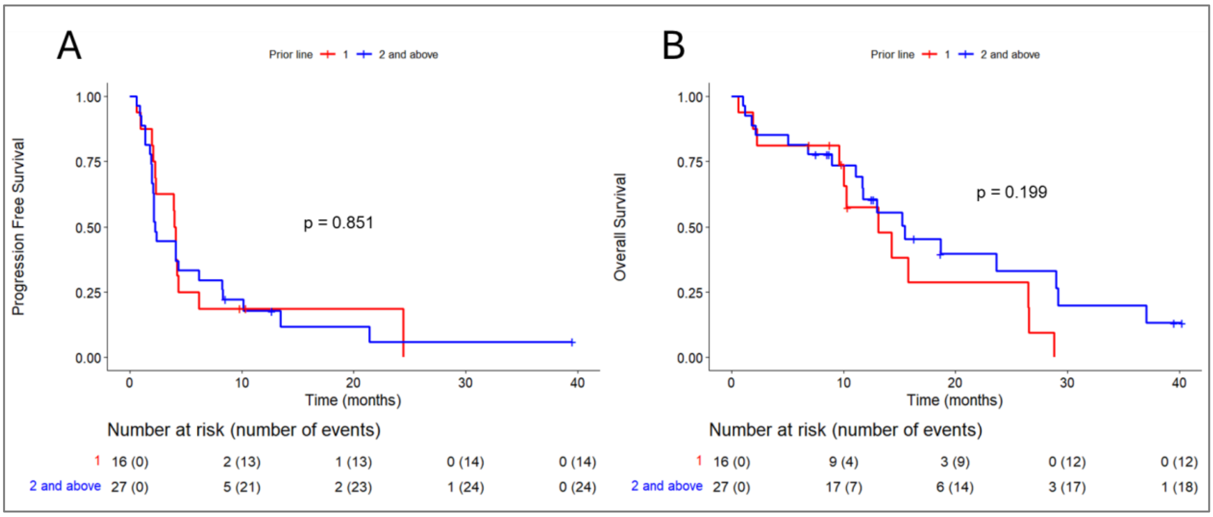


Supp. Fig 4. Kaplan-Meier estimates of (A) progression free survival and (B) overall survival according to the number of prior lines of therapy.
